# Knowledge transfer to enhance the performance of deep learning models for automated classification of B-cell neoplasms

**DOI:** 10.1101/2021.03.03.21252824

**Authors:** Nanditha Mallesh, Max Zhao, Lisa Meintker, Alexander Höllein, Franz Elsner, Hannes Lüling, Torsten Haferlach, Wolfgang Kern, Jörg Westermann, Peter Brossart, Stefan W. Krause, Peter M. Krawitz

## Abstract

Multi-parameter flow cytometry (MFC) is a cornerstone in clinical decision making for hematological disorders such as leukemia or lymphoma. MFC data analysis requires trained experts to manually gate cell populations of interest, which is time-consuming and subjective. Manual gating is often limited to a two-dimensional space. In recent years, deep learning models have been developed to analyze the data in high-dimensional space and are highly accurate. Such models have been used successfully in histology, cytopathology, image flow cytometry, and conventional MFC analysis. However, current AI models used for subtype classification based on MFC data are limited to the antibody (flow cytometry) panel they were trained on. Thus, a key challenge in deploying AI models into routine diagnostics is the robustness and adaptability of such models. In this study, we present a workflow to extend our previous model to four additional MFC panels. We employ knowledge transfer to adapt the model to smaller data sets. We trained models for each of the data sets by transferring the features learned from our base model. With our workflow, we could increase the model’s overall performance and more prominently, increase the learning rate for very small training sizes.

## Introduction

Multi-parameter flow cytometry (MFC) is a powerful and high-throughput technique that allows for rapid quantification of markers on cells in suspension^1^. Today, it is a critical step in both research and clinical decision making for leukemia^2,3^ and other hematological diseases. However, a significant limitation of the method is that data are analyzed manually, including gating cell populations of interest in a two-dimensional scatter plot, which is time-consuming and subjective^4,5^. In recent years, more advanced computation methods involving deep learning have become available that can accurately classify disease subtypes based on cell type identification from cytological images^6^ and perform automated classification of MFC data into diagnosis labels^7,8^. However, such models used for MFC analysis are limited to the MFC panel they are trained on and do not produce the same performance on a different MFC panel.

The flow cytometry panel design across various laboratories varies depending on the markers to be analyzed and the cytometer available. In many cases, the number of markers needed to be analyzed exceeds the number that the cytometer can measure in a single run. Standard practice is to aliquot a sample into multiple tubes, each of which often includes a set of shared or backbone markers^9^. This process is standard for modern clinical diagnostic of MFC data; especially, when immunophenotyping leukemia and lymphoma. Furthermore, the choice of markers depends on the diagnostic workflow and is not standardized. These differences result in different antibody (MFC) panels being used in different laboratories. Thus, AI models must be robust and adapt to different MFC panels across laboratories and within the same laboratory when the diagnostic panel is modified due to transition to a new cytometer.

This study extends our previous model^8^, where we trained an AI model to classify seven B-cell neoplasm subtypes plus healthy controls for a 9-color panel to work with multiple MFC panels by employing transfer learning. Transfer learning (TL) is a technique to improve the performance of a new task by transferring knowledge from a related task that has already been learned^10^. In our case, both the base and target tasks to be learned are the same: classification of B-cell neoplasms into diagnosis labels. However, the MFC protocol with which a sample is acquired is subject to inter-laboratory variability and changes over time in terms of the number of tubes per sample, markers measured, marker-fluorochromes conjugates and other protocol parameters.

To handle the differences across MFC panels and achieve maximum knowledge transfer, we merge FCS data from individual tubes of a sample into a single combined FCS file using the nearest neighbor (NN) method. This method assumes that a cell in one tube is identical to its NN in another tube in terms of the shared markers and can thus be used to impute missing marker values^11,12^. The expression vectors of all the NNs across tubes are merged, creating a single, high-dimension matrix of cellular expression across all tubes. Nearest neighbor merging has proven effective as part of classification pipelines^9,13^, while other merging methods are better suited for deep profiling^14^. We use NN merge in conjunction with transfer learning to generalize our model and achieve a higher learning rate with fewer training samples.

## Methods

### Flow cytometry data

The base data set^8^ consists of around 18,000 training samples acquired using a 9-color MFC panel at Munich Leukemia Laboratory (MLL) between 2017-2018. Four additional MFC target data sets were acquired with different MFC panel compositions. The number of samples per cohort in each of the four target data sets are summarized in Table 1.

**Table 1.**
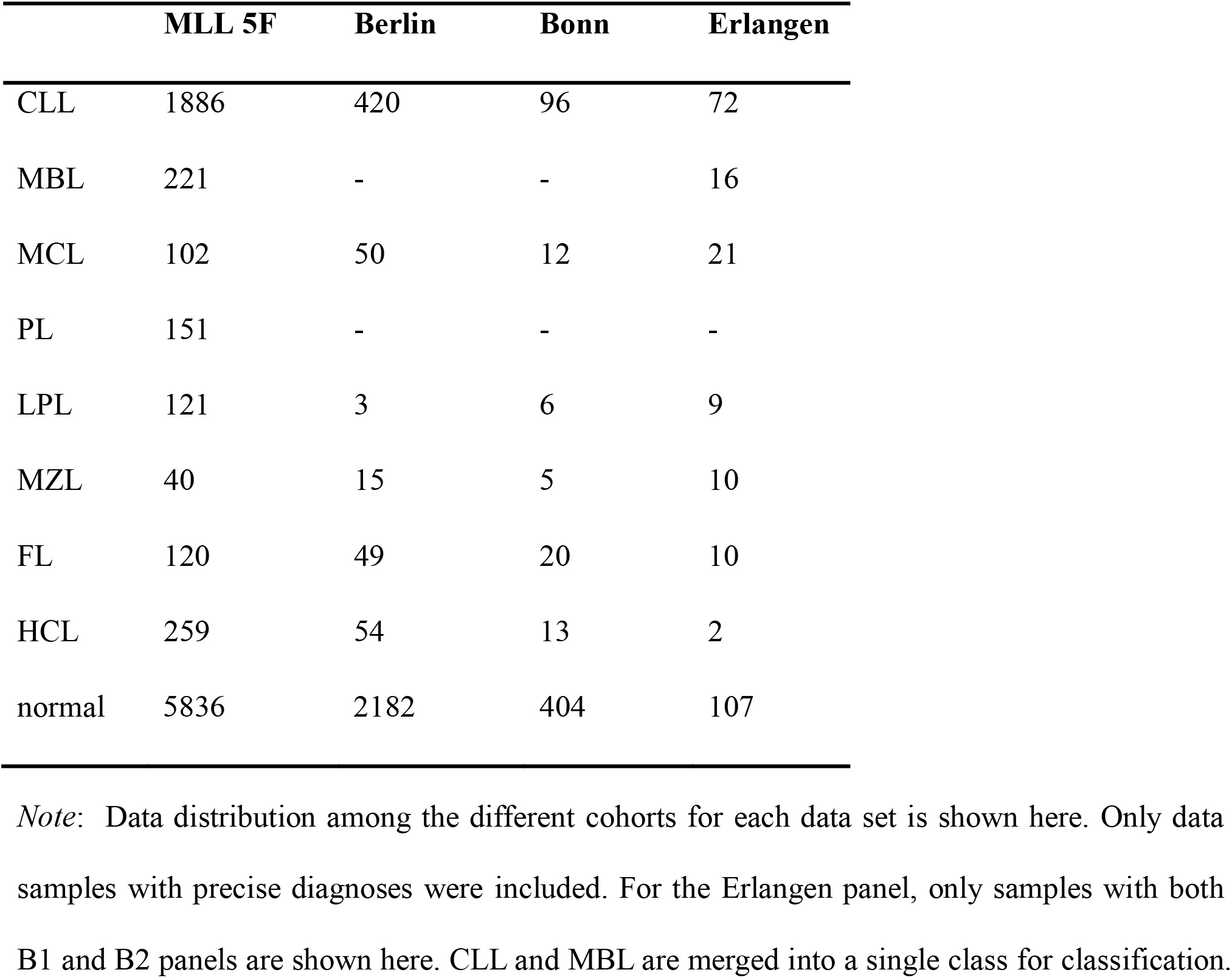
B-cell lymphoma cohorts in target data sets

#### Munich 5-color panel

A 5-color panel consisting of 10,079 samples was acquired at MLL between January 01, 2011, and December 31, 2012. For the assessment of B-cell neoplasms (B-NHL), a panel consisting of seven 5-color combinations of monoclonal antibodies was used in all samples to analyze the surface expression of 20 antigens. A detailed antibody-color combination is given in the Supporting Information Table S1a. In the following, we refer to this panel as the MLL5F panel.

#### Bonn 9-color panel

The second data set was obtained from the University Hospital Bonn, consisting of 525 samples measured between January 01, 2018, and December 31, 2019. For B-NHL assessment, a panel consisting of two 9-color combinations of monoclonal antibodies was used to analyze 16 antigens’ surface expression. Detailed MFC panel information is given in the Supporting Information Table S1b. In the following, we refer to this panel as the Bonn panel.

#### Berlin 8-color panel

For the third data set, an 8-color panel consisting of 2,773 routine diagnostic samples from patients with suspected B-cell neoplasms analyzed between January 01, 2016, and December 31, 2018, was obtained from the Berlin Hematology laboratory. The B-NHL assessment panel consisted of four 8-color combinations of monoclonal antibodies. Supporting Information Table S1c details the MFC panel used. In the following, we refer to this panel as the Berlin panel.

#### Erlangen panel

A fourth target data set was obtained from the University Hospital Erlangen. The data set consisted of 1,626 routine diagnostic samples from patients with suspected B-NHL analyzed between January 01, 2014, and July 31, 2020. For the assessment of B-NHL, a screening panel (B1) consisting of one 10-color combination of monoclonal antibodies was used to analyze the surface expression of 9 antigens. Next, a secondary panel (B2) to identify the B-NHL subtype was used where necessary. For the identification of HCL, a third panel (B3) was used. All three panels are described in detail in Supporting Information Table S1d. For this study, we only consider the 247 samples with both B1 and B2 panels.

All samples were analyzed on Navios cytometers (Beckman Coulter, Miami, FL). All diagnoses were verified with additional tests from histology, cytomorphology, in-situ fluorescence hybridization (FISH), and only cases with unambiguous labels were used to train the models. Further, only samples obtained from peripheral blood or bone marrow aspirate were included in the analysis. Flow cytometry data are stored in the FCS 2.0 format^15^. The compensated FCS 2.0 data segment has been used in the analysis.

### Transfer learning

An overview of the process is shown in Fig 1. Before knowledge transfer, we merge multiple aliquots (tubes) per sample into a single large FCS data file. In our previously published model, we processed individual tubes of each sample separately, resulting in a CNN architecture that depended on the number of tubes per sample. Without the initial merge step, such a network’s transferability between data sets with a different number of tubes per sample is very low - we can only transfer knowledge from the dense layers (Supplementary Fig 1). Merging multiple aliquots allows for maximum transfer between the networks - weights from all layers can now be transferred.

**Figure 1.**
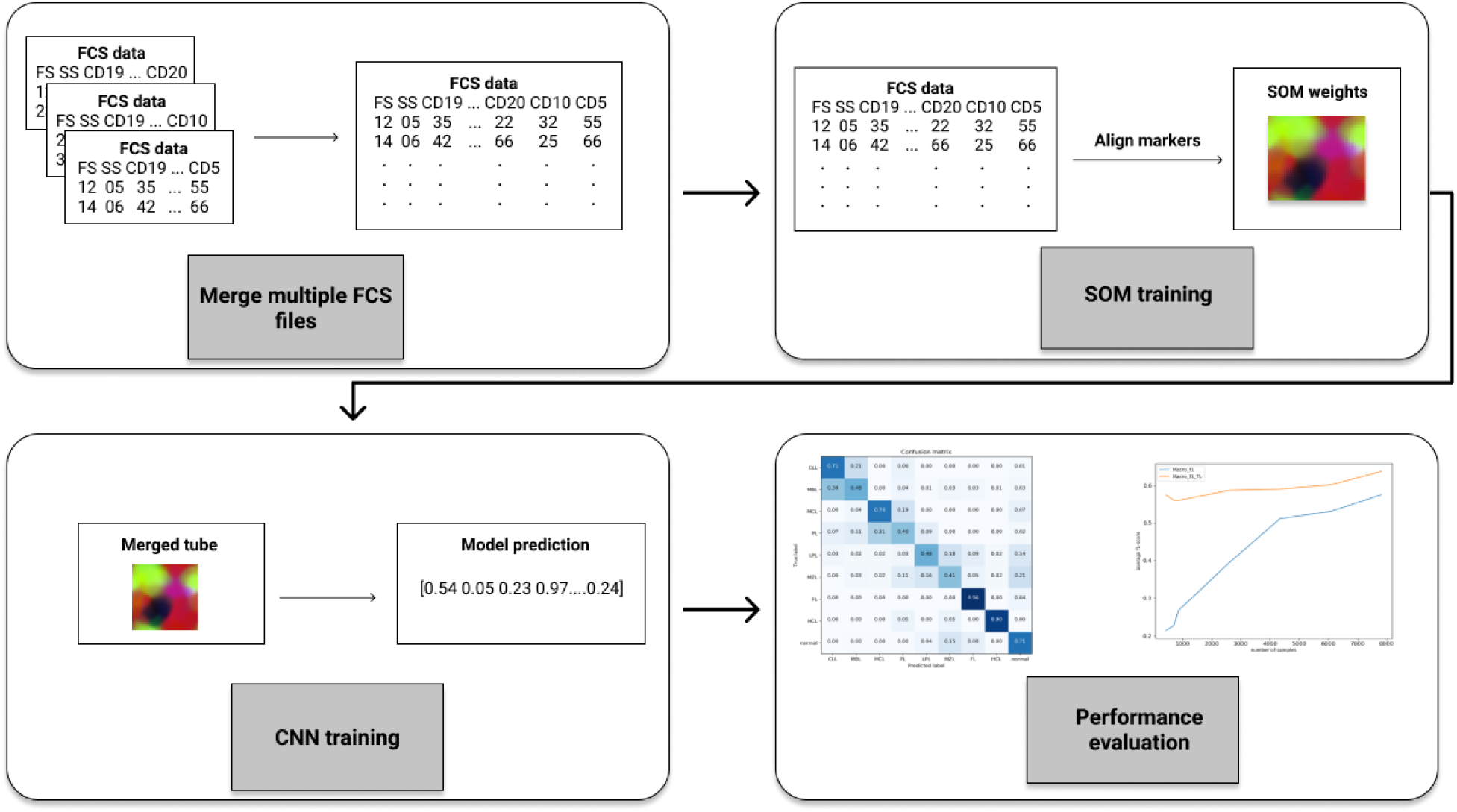
Overview of the knowledge transfer pipeline. Multiple FCS files per sample are merged using the NN merge. Individual 2D SOMs are then generated using the merged FCS file for each sample. The weights of the SOM nodes are used as input for a CNN that predicts lymphoma subtypes. For each data set, two models – standalone and model with knowledge transfer are generated. The gain from knowledge transfer is then evaluated by comparing the performance of both models.

### Merge

The merge process is depicted in Fig 2. The process for matching events between different data files is as follows.

**Figure 2.**
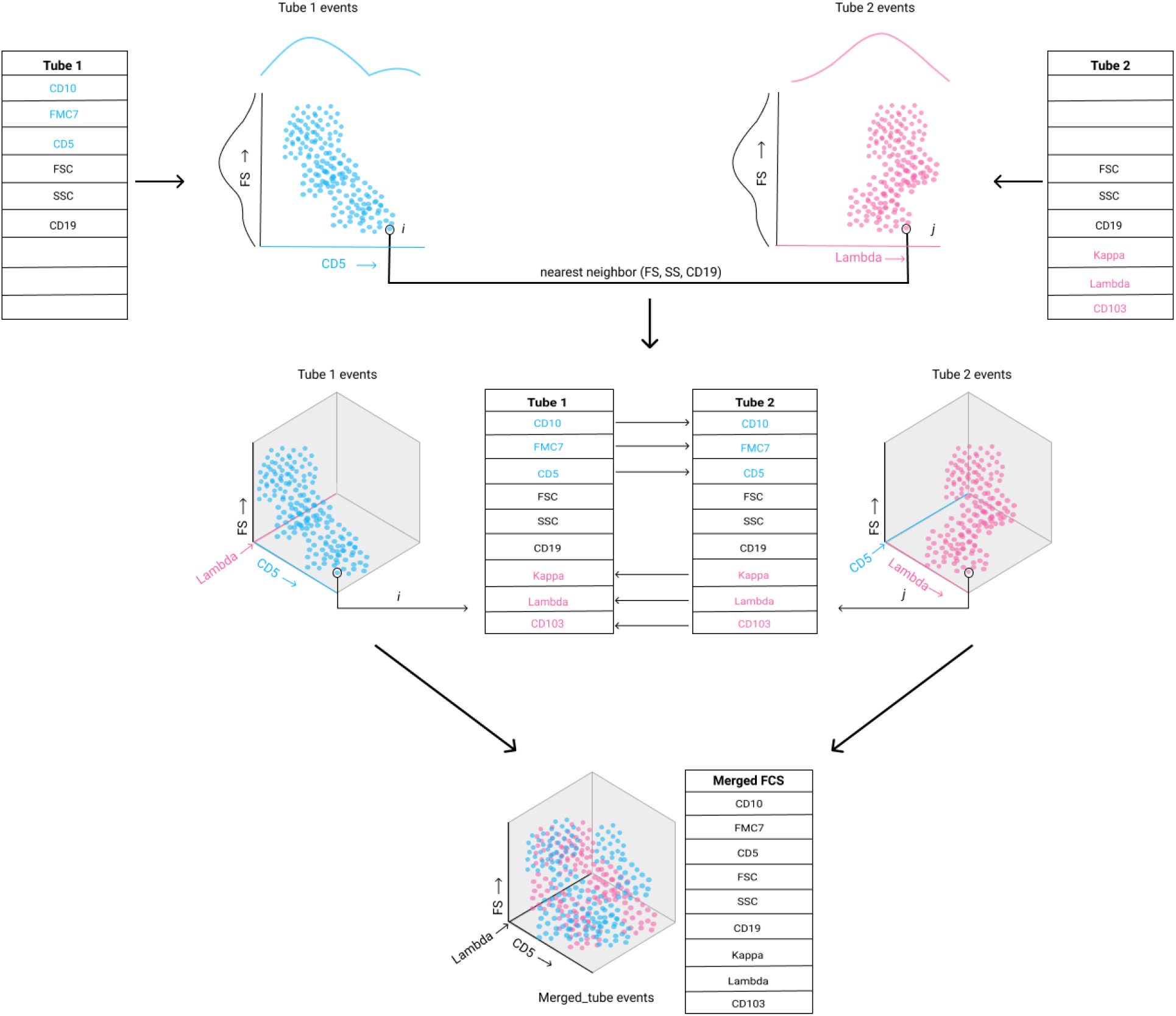
Merge overview. Overview of NN merge is shown here for two tubes with three shared markers. Each tube has three tube-specific markers: CD10, FMC7, and CD5 are tube 1 specific markers, while tube 2 specific markers are Kappa, Lambda, and CD103. Events are shown in a two-dimensional space with one common marker (FS) and one tube-specific marker (CD5 for tube 1 and Lambda for tube 2). For each event “*i”* in tube 1, the nearest neighbor in tube 2, “*j*,” is computed in terms of the shared markers (FS, SS, CD19). Next, tube 2 specific markers from “*j*” are copied over to “*i*”; tube1 specific markers from “*i*” are copied over to “*j*.” The process is repeated for all the events in tube1 and 2. All the tube 1 events will have imputed values for tube 2 specific markers (Kappa, Lambda, and CD103) and tube 2 events will have imputed tube 1 specific markers (CD5, CD10, and FMC7); these events can now be analyzed for the imputed markers that were previously missing. Finally, the expression vectors of all events across tubes are merged, resulting in a combined FCS file consisting of events from both tubes with all the measured parameters.

*Step1:* Determine the common markers for each of the data sets (Supplementary Information Table S2). The common markers are used as the vector to calculate the distance between events in different data files.

*Step2:* Take tube *i* (start with the first tube; *i*=1), and iterate over all of the remaining tubes *j*

*Step3:* For each event in tube *i*, calculate the nearest neighbor (NN) in tubes *j*

*Step4:* Copy the tube specific marker values from the computed NNs in tube *j* to the events in tube *i*

*Step5:* Increment *i*, repeat the above steps. Events in each tube will now have imputed values for markers that were measured in a different tube

*Step6:* Merge all the events across all tubes into a single large matrix

The resulting data file obtained after merging the original data files and calculating each event’s values was a file containing information about all parameters measured in all multicolor staining for each of the events recorded. For the MLL5F panel, we merge tubes 2, 3, 4, 5, and 7. Thus, each merged/calculated data file contained information about all 18 parameters measured for each of the ≥2.5 × 10^5 events analyzed per sample (five aliquots/sample × ≥5 × 10^4 events/ aliquot). The tubes merged for the different data sets and the merge parameters are described in Supplementary Information Table S2.

We implement the merge using scikit-learn API^16^. The code used for merging files is available at https://github.com/NandithaMallesh/fcsmerge.git.

### Self-Organizing Map (SOM)

A SOM is a network of interconnected nodes, ordered in a two-dimensional topology, which can be used for unsupervised clustering of high dimensional data^17^. SOMs were used as a method to reduce the dimensionality of the data while preserving its spatial structure. We first generate individual SOM for each of the merged FCS samples. Each node in the SOM is associated with a “weight” vector representing the FCS data. Individual SOM transformation uses pre-initialized node weights from a reference SOM. Reference SOM from the base model is used as the pre-initialized weights for the target data sets to ensure the same initial tree structure. Markers are aligned to the base data set by matching FS, SS, and as many CD markers as possible by disregarding the associated fluorochromes. In case of missing markers in the target sets, they are set to “n/a,” any new CD markers in the target set that are not found in the base set are ignored. The SOM implementation was adapted to account for missing data values by modifying the training process^18^. We introduce a masking matrix with values in {0, 1} for each value in the original data. “1” indicates that the data value is valid, and “0” indicates the data is invalid, and the data point should be ignored for any calculations. The SOM training is then adjusted to use the mask values to ignore invalid data points for the best-matching unit (BMU) calculation and weight updates.

### Prediction model

The modified CNN architecture for the merged data is shown in Supplementary Fig 3. The model generates predictions using SOM node weights for a number of classes. The weights are first processed with three convolution layers with decreasing filter sizes, followed by a global max-pooling layer that summarizes filters across the SOM map’s spatial dimension. Next, two dense layers combine information, and a final softmax layer generates class predictions. Models are trained using Adam optimizer^19^ with a learning rate of 0.001. A global weight decay of 5e-6 was applied to all layers. The model is implemented in Keras^20^. We trained a base model for the merged base data set with the modified CNN architecture.

We trained two models for each target data set: a standalone model without knowledge transfer and a second model with knowledge from the base model. The weights for each layer in the target network are initialized with trained weights from the base model’s corresponding layer. The standalone models’ hyperparameters are kept identical to the base model - 20 epochs, learning rate of 0.001, and a global decay of 5e-6. For the second set of models with transfer learning, we used the same learning rate and global decay while the number of epochs was reduced to 15. Further, the two dense layers are frozen by setting the “trainable” hyperparameter as false. When using transfer learning, the norm is to freeze the convolution layers to avoid overfitting. However, in our case, the MFC panel composition is different from the base data. To account for changes in the panel, we keep the convolution layers unfrozen and retrain them to learn the filters for the target MFC panel. We freeze the two dense layers that combine information for generating class prediction since the classes to be predicted are the same as in the base task.

We perform 10-fold validation for all the target models to avoid any bias resulting from a single random train-test split, especially for the smaller data sets. Models are trained on the training split, and performance metrics are calculated on the validation split. The average scores across 10-folds are reported as the final performance measure.

### Performance metrics

Precision and recall per group were defined on the true label of each case. Prediction performance was evaluated with average-per-group F1 scores calculated as avg 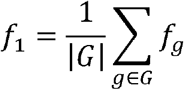, with 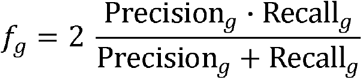 Where *G* is the set of all groups.

Weighted F1 score was calculated as the group-size weighted average of per-group F1 scores weighted 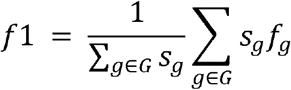 with *s*_*g*_ as the number of cases in Group *g*.

For each model, we calculate top-1-accuracy rate of the classifier for the eight classes: chronic lymphocytic leukemia and its predecessor monoclonal B-cell lymphocytosis (CLL/MBL), marginal zone lymphoma (MZL), mantle cell lymphoma (MCL), prolymphocytic leukemia (PL), follicular lymphoma (FL), hairy cell leukemia (HCL), and lymphoplasmacytic lymphoma (LPL) and healthy controls. As MBL is defined by fewer than 5,000 cells/ul in peripheral blood with the typical CLL profile in flow cytometry, we listed MBL as a separate class in the learning curves and ROC curves for a fine-grained analysis of classification sensitivity.

## Results

### Comparing merged and original data sets

To evaluate the quality of the merged data set, we use Jensen-Shannon distance to quantify the similarity between the distributions of markers in the original and merged data sets, resulting in values between zero (identical distributions) to one (totally disjoint distributions). If *p* and *q* are the probability distributions of a marker in the original and merged data, then the Jensen-Shannon distance (JSD) is calculated as the square root of Jensen-Shannon divergence^21^:

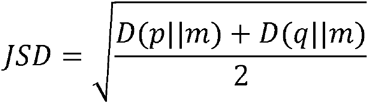

Where m is the pointwise mean of *p* and *q*, and D is the Kullback-Leibler divergence^22^.

We compute the JSD for each of the non-shared markers between the original and merged sample for all data sets. We obtained a mean JSD score of less than 0.1 across all the markers for all the data sets, indicating good agreement between the merged and original data set in terms of marker distribution. The average JSD across all markers for each data set and the individual JSD score for every marker is reported in Supplementary Fig 2. Although JSD scores do not evaluate the extent to which a cell population (based on co-expression of markers) is preserved in the merged data, the scores provide a way of evaluating how the imputation affected individual marker distribution. Further, the original and merged models’ performance for the base data was compared to evaluate the effect of NN merge on the CNN classification. The merged base model achieved an overall weighted F1 score of 0.94 and an average F1 score of 0.74. In comparison, the original model trained with the unmerged FCS data from tubes 1 and 2 achieved an overall weighted F1 score of 0.94 and an average F1 score of 0.75.

### Evaluation of knowledge transfer

To evaluate knowledge transfer, we compare the performance of the target models with and without transfer learning. A 10-fold validation is performed on both the standalone and transfer learning models for each target data set. For each model, weighted and average F1 scores are calculated. The models with transfer learning show a significant improvement in F1 scores, especially the average F1 scores for all the data sets (Fig 3). The delta in the performance between the data sets may be attributed to the size of the data set, quality of the original data, and the quality of the merged data with imputed marker values. The effect of the quality of merged data on the classification score is discussed in Supplementary Fig 2d. The overall scores obtained by averaging the F1 scores over 10-folds and the 95% CI values for the four data sets are reported in Table 2. The ROC curves and mean AUC for standalone and transfer learning models are shown in Supplementary Fig 4.

**Table 2.** Performance metrics

|  | **MLL 5F** | **Berlin** | **Bonn** | **Erlangen** |
| --- | --- | --- | --- | --- |
| With_TL | f1_weighted: 0.93<br>95% CI: (0.92,<br>0.93) | f1_weighted: 0.93<br>95% CI: (0.91,<br>0.95) | f1_weighted: 0.85<br>95% CI: (0.81,<br>0.88) | f1_weighted: 0.80<br>95% CI: (0.73,<br>0.87) |
|  | f1_avg: 0.64<br>95% CI: (0.61,<br>0.66) | f1_avg: 0.62<br>95% CI: (0.54,<br>0.71) | f1_avg: 0.50<br>95% CI: (0.41,<br>0.59) | f1_avg: 0.52<br>95% CI: (0.40,<br>0.64) |
| Standalone | f1_weighted: 0.92<br>95% CI: (0.91,<br>0.93) | f1_weighted: 0.92<br>95% CI: (0.90,<br>0.93) | f1_weighted: 0.76<br>95% CI: (0.69,<br>0.83) | f1_weighted: 0.69<br>95% CI: (0.63,<br>0.74) |
|  | f1_avg: 0.57<br>95% CI: (0.54,<br>0.59) | f1_avg: 0.52<br>95% CI: (0.45,<br>0.59) | f1_avg: 0.40<br>95% CI: (0.26,<br>0.53) | f1_avg: 0.35<br>95% CI: (0.31,<br>0.40) |

*Note:* Weighted and Average F1 score along with 95% CI values for the four target data sets for models with knowledge transfer and standalone models without transfer learning. The F-scores were calculated as an average of the 10-fold scores for each data set.

**Figure 3.**
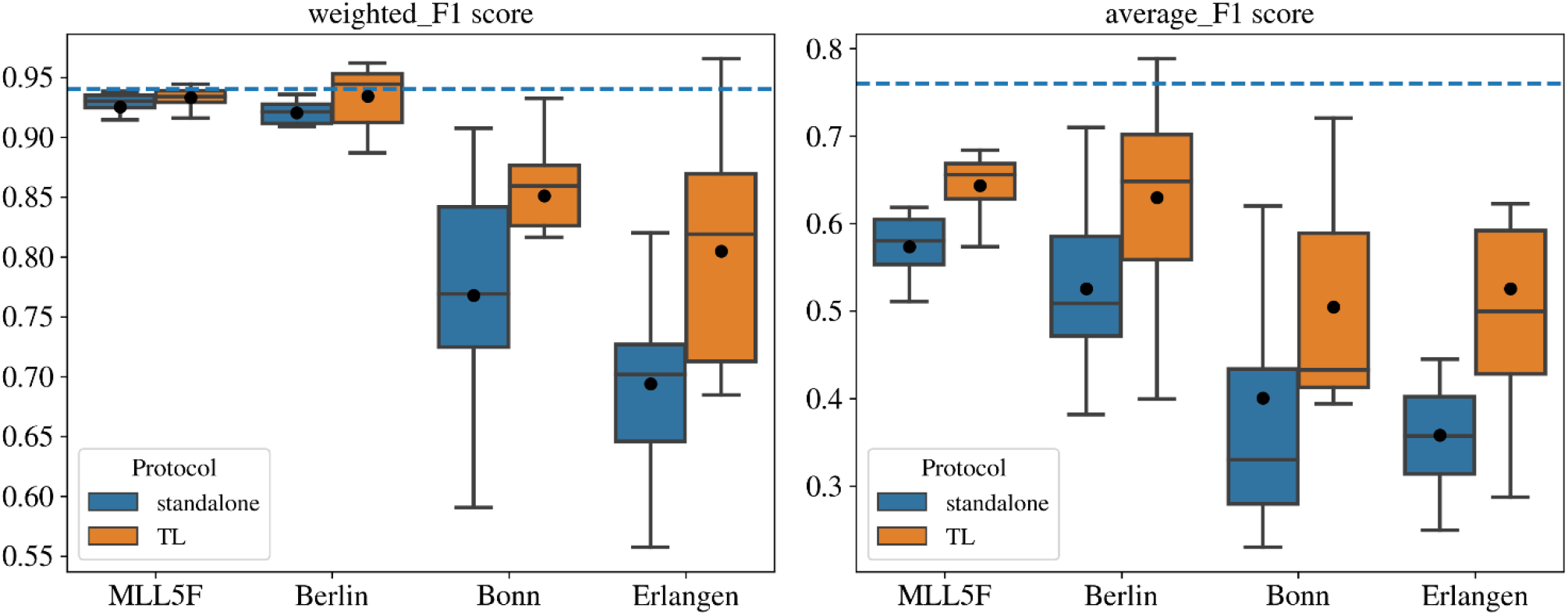
Performance for Standalone Vs. Transfer learning. The Box plots show F1 scores along with mean values obtained: on the left, weighted_F1_scores are plotted for each data set, and on the right, the avergae_F1_scores are shown. The blue dotted line across the plots represents the previously reported base model’s performance, which is considered as the expert-level accuracy for this work. The transfer learning models perform better in all four data sets. These models achieve a higher F1 score, especially the average f1 score. A significant increase in average_F1 score is seen for MLL5F (p-value = 1.805e-03) and Erlangen (p-value = 3.194e-02) panels. For Bonn and Berlin panels, we achieved a p-value of 6.838e-01 and 1.659e-01, respectively. All p-values were computed using an independent t-test with Bonferroni correction.

### Learning curve analysis

Here, we describe two use cases that change the MFC diagnostic panel and require an AI model to be adapted. We use our current workflow to adapt the base model for both cases and analyze the model’s learning curves for each case. Transfer learning with merge increases the models’ overall performance and the models have a higher start on the learning curve for smaller sample sizes.

#### Case 1: Transition to a new cytometer within the same lab

In multicolor flow cytometry (MFC) diagnostics, switching to a device that supports more fluorochromes per measurement is a common transition in a diagnostic laboratory that optimizes its workflows by updating its equipment. Usually, this process involves a few weeks where samples are measured with both protocols, the old one validating the new one. However, this means that only a few samples from the new protocol are available to train the new classifier. By using knowledge transfer, we show transition can be handled quickly by adapting the existing AI model. We set up a transition scenario from a 5-color cytometer to 9-color using our MLL5F and MLL9F data sets. We train a model with the MLL5F panel and use this as the base network to train a new model for the MLL9F panel. The training set for the target network included a varying number of samples per group ranging from 5 to 50. F1 scores were recorded for each iteration. The learning curve (Fig 4a) with TL shows a higher start and asymptote for the target network, the confusion matrix obtained with five training samples per class shows a significant improvement in classification, especially for the smaller classes.

**Figure 4.**
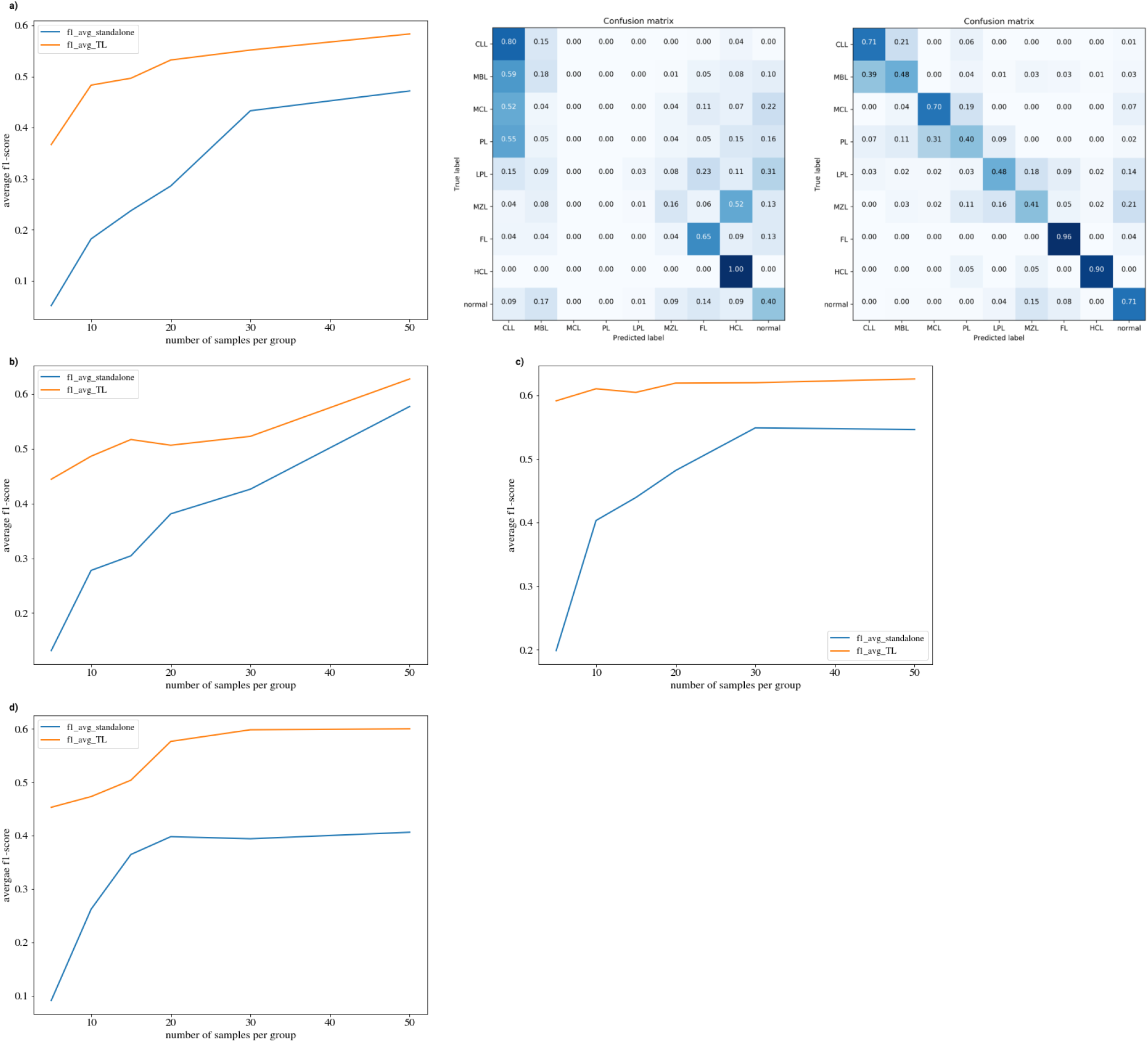
Experiments: a) The learning curve obtained for average f1 scores for the transition experiment is shown on the left. The model with transfer learning has a significantly higher score for small training size. The confusion matrices for the standalone model and model_with_TL with five training samples per class are shown on the right. As seen in the confusion matrix, with knowledge transfer, the classification of rare subtypes improves. The learning curves for inter-laboratory adaptation for Bonn, Berlin, and the Erlangen panel is shown in b), c), and d). The learning curves were obtained for training sizes ranging from 5 samples to 50 samples per class. For instances where the total number of available samples for a class is less than 50, we resample the existing data to create sufficient training samples for each class. The f1 curves show the same trend as in the transition experiment – higher start and asymptote with transfer learning and an overall performance enhancement.

#### Case 2a: Model adaptability across laboratories

MFC diagnostic workflows are relatively similar across laboratories. However, the MFC panel used for diagnosis varies depending on the cytometer and antibodies measured. For an AI model, the reported performance is valid for the given MFC panel. When the model is used to interpret different MFC data, the performance drops significantly without changes to the underlying architecture and parameters. Training a new model with the new data requires longer training time and large data sets. Here, we demonstrate our workflow to extend a model trained on a specific MFC panel with an extensive training data set to different MFC panels with lesser data.

We use our merged base model to train new models for Bonn and Berlin panels. Both target models show a significant increase in overall performance with transfer learning. The target models are trained with an increasing number of samples per group ranging from 5 to 50. The learning curves for F1 scores show a significantly higher start and overall performance in cross laboratory adaptation with our workflow (Fig 4b and 4c).

#### Case 2b: Cross laboratory adaptation with different diagnostic setting

For the Erlangen data set, a screening panel is used to diagnose B-cell neoplasms with a separate classification panel for further subtype determination. We trained a model with the same architecture and parameters as our base model for the screening panel to obtain a “normal” vs. “pathological” binary classification. The resulting model could classify 86% of pathological samples and 96% of normal samples correctly. Further, we used our current workflow for the 247 samples with both screening and the classification panel (B1 and B2). Here, we employ knowledge transfer as described and see an overall gain in the average F1 score from 0.33 to 0.52. The learning curve (Fig 4d) shows a higher start and asymptote similar to the other three data sets.

## Discussion

Our proposed workflow combines knowledge transfer with FCS data merging that enables existing models to adapt to various MFC panels. We extend our base model to four additional data sets with a varying number of tubes per sample and markers with no changes to the model architecture and training parameters. Here, we show knowledge transfer enhances the overall performance for new models by allowing for already learned features from a large data set to be transferred to smaller data sets.

Merging multiple aliquots is a known approach for increasing computational depth for deep phenotyping and FCS analysis^23^. While NN merge is known to sometimes introduce a spurious combination of markers into the imputation results^24^, this did not lead to a reduced performance of our classification model. In our case, merging multiple aliquots increases the convolution neural network’s feature space by combining markers measured in different tubes. It also allows us to maximize our networks’ transferability that is essential for a successful knowledge transfer.

With our workflow, the target models achieve an overall performance close to the previously reported expert-level accuracy^8^. For the Berlin panel, the transfer learning model achieved a median weighted F1 score of 0.94, the same as expert-level performance, which was otherwise not possible. The learning rate with knowledge transfer is also much higher; the models achieve significantly higher performance for very small training sizes. In the context of transition to a new cytometer, this would allow an already deployed AI model to be quickly adapted to the new protocol without having to wait for a considerable time for enough samples to become available for the new protocol.

For our knowledge transfer, we align FCS channels by matching CD markers while ignoring the fluorochromes. While any missing markers are handled efficiently, the current workflow will ignore new markers. The performance may be improved further with partial knowledge transfer techniques, where features from existing channels are transferred while the model is trained to learn the new channels present in the new protocol^25^.

In conclusion, we present a workflow to extend deep learning models to multiple MFC panels and achieve high accuracy for the multi-label classification across data sets. Here, we address some of the previous challenges for automated flow cytometry classification by allowing models to be trained with smaller training sizes and generalizing models to work with multiple MFC panels. Our workflow is a step towards making deep learning models robust so that AI for diagnostic MFC can move from the “proof of concept” stage into routine diagnostics.

## Data Availability

All data are available upon reasonable request. Please send a request to Peter Krawitz at

## Acknowledgments

This study was supported by the German Research Council (Deutsche Forschungsgemeinschaft, DFG), by project 315041274, awarded to P.M.K.

## Ethics Approval

IRB or ethics approval does not apply as the study was conducted on fully anonymized retrospective patient data. A waiver was granted by the University of Bonn Medical Faculty Ethics Committee.

## Code Availability

The source code used for merging FCS files, model generation, and transfer learning is available under an open source license from the following git repositories: https://github.com/NandithaMallesh/mergeTL and https://github.com/NandithaMallesh/fcsmerge. All visualizations were implemented in matplotlib.

## Data Availability

All data are available upon reasonable request. Please send a request to Peter Krawitz at

## Bibliography

1. Shapiro, H. M. Practical flow cytometry. (Wiley-Liss, 2003).

2. Henel, G. & Schmitz, J. L. Basic theory and clinical applications of flow cytometry. Laboratory Medicine vol. 38 (2007).

3. Craig, F. E. & Foon, K. A. Flow cytometric immunophenotyping for hematologic neoplasms. Blood vol. 111 (2008).

4. Bendall, S. C. & Nolan, G. P. From single cells to deep phenotypes in cancer. Nature Biotechnology vol. 30 (2012).

5. O’Neill, K., Aghaeepour, N., Špidlen, J. & Brinkman, R. Flow Cytometry Bioinformatics. PLoS Comput. Biol. 9, (2013).

6. Matek, C., Schwarz, S., Spiekermann, K. & Marr, C. Human-level recognition of blast cells in acute myeloid leukaemia with convolutional neural networks. Nat. Mach. Intell. 1, (2019).

7. Ng, D. P. & Zuromski, L. M. Augmented Human Intelligence and Automated Diagnosis in Flow Cytometry for Hematologic Malignancies. Am. J. Clin. Pathol. (2020) doi:10.1093/ajcp/aqaa166.

8. Zhao, M. et al.. Hematologist-Level Classification of Mature B-Cell Neoplasm Using Deep Learning on Multiparameter Flow Cytometry Data. Cytom. Part A 97, (2020).

9. Van Dongen, J. J. M. et al.. EuroFlow antibody panels for standardized n-dimensional flow cytometric immunophenotyping of normal, reactive and malignant leukocytes. Leukemia 26, (2012).

10. Weiss, K., Khoshgoftaar, T. M. & Wang, D. D. A survey of transfer learning. J. Big Data 3, (2016).

11. Pedreira, C. E. et al.. Generation of flow cytometry data files with a potentially infinite number of dimensions. Cytom. Part A 73, (2008).

12. Abdelaal, T. et al.. CyTOFmerge: Integrating mass cytometry data across multiple panels. Bioinformatics 35, (2019).

13. Costa, E. S. et al.. Automated pattern-guided principal component analysis vs expert-based immunophenotypic classification of B-cell chronic lymphoproliferative disorders: A step forward in the standardization of clinical immunophenotyping. Leukemia 24, (2010).

14. O’Neill, K. et al.. Deep profiling of multitube flow cytometry data. Bioinformatics 31, (2015).

15. Dean, P. N., Bagwell, C. B., Lindmo, T., Murphy, R. F. & Salzman, G. C. Introduction to flow cytometry data file standard. Cytometry 11, (1990).

16. Buitinck, L. et al.. {API} design for machine learning software: experiences from the scikit-learn project. in ECML PKDD Workshop: Languages for Data Mining and Machine Learning 108–122 (2013).

17. Kohonen, T. The Self-Organizing Map. Proc. IEEE 78, (1990).

18. Samad, T. & Harp, S. A. Self-organization with partial data. Netw. Comput. Neural Syst. 3, (1992).

19. Kingma, D. P. & Ba, J. L. Adam: A method for stochastic optimization. in 3rd International Conference on Learning Representations, ICLR 2015 - Conference Track Proceedings (2015).

20. Chollet, F. Keras. J. Chem. Inf. Model. 53, (2013).

21. Naghshvar, M., Javidi, T. & Wigger, M. Extrinsic Jensen–Shannon Divergence: Applications to Variable-Length Coding. IEEE Trans. Inf. Theory 61, 2148–2164 (2015).

22. Kullback, S. & Leibler, R. A. On Information and Sufficiency. Ann. Math. Stat. 22, 79–86 (1951).

23. Robinson, J. P., Durack, G. & Kelley, S. An innovation in flow cytometry data collection and analysis producing a correlated multiple sample analysis in a single file. Cytometry 12, (1991).

24. Lee, G., Finn, W. & Scott, C. Statistical file matching of flow cytometry data. J. Biomed. Inform. 44, (2011).

25. Hassan, A. Transfer Learning from RGB to Multi-band Imagery. Azavea https://www.azavea.com/blog/2019/08/30/transfer-learning-from-rgb-to-multi-band-imagery/ (2019).

