## Supplement Data for "Knowledge transfer to enhance the performance of deep learning models for automated classification of B-cell neoplasms"

**Supplementary Material**

|  | **FITC** | **PE** | **ECD** | **PC5.5** | **PC7** |
| --- | --- | --- | --- | --- | --- |
| Tube 1 | IgG1a | IgG1a | IgG1a | IgG1a | CD45 |
| Tube 2 | CD79b | CD5 | CD19 | CD20 | CD45 |
| Tube 3 | FMC7 | IgM | CD19 | CD10 | CD45 |
| Tube 4 | CD103 | CD23 | CD19 | CD22 | CD45 |
| Tube 5 | kappa | lambda | CD19 | CD38 | CD45 |
| Tube 6 | CD8 | CD4 | CD3 | CD56 | CD45 |
| Tube 7 | - | CD11c | CD19 | CD25 | CD45 |

*Supplementary Table S1a*. Antibody color combinations used in the 5-color MLL5F panel. Colors are shown in the header row, and individual antibodies are shown for each tube. Further, forward scatter (FS) and side scatter (SS) were measured in all tubes. Tube 1 is used for isotope control and is not considered for FCS merging, SOM generation, and classification.

|  | **FITC** | **PE** | **ECD** | **PC5.5** | **PC7** | **APC** | **AA700** | | **AA750** | **PB** |
| --- | --- | --- | --- | --- | --- | --- | --- | --- | --- | --- |
| Tube 1 | FMC7 | CD23 | CD19 | CD11c | CD200 | CD79b | | CD5 | CD43 | CD20 |
| Tube 2 | Kappa | Lambda | CD19 | CD10 | CD22 | CD103 | | CD25 | CD38 | CD20 |

*Supplementary Table S1b*. Antibody color combinations used in the 9-color Bonn panel. Forward scatter (FS) and side scatter (SS) were measured in all tubes.

|  | **FITC** | **PE** | **ECD** | **PC5.5** | **PC7** | **APC** | **PB** | **KrOr** |
| --- | --- | --- | --- | --- | --- | --- | --- | --- |
| Tube 1 | IgG | IgG | IgG | IgG | IgG | IgG | IgG | CD45 |
| Tube 2 | Kappa | Lambda | CD19 | CD5 | CD38 | CD10 | CD20 | CD45 |
| Tube 3 | FMC7 | CD23 | CD19 | CD3 | - | CD79 | CD22 | CD45 |
| Tube 4 | CD43 | IgM | CD19 | CD25 | CD11C | CD103 | CD5 | CD45 |

*Supplementary Table S1c*. Antibody color combinations used in the 8-color Berlin panel. Tube 1 is used as isotope control and is not considered in the workflow.

|  | **FITC** | **PE** | **ECD** | **PC5.5** | **PC7** | **APC** | **APC750** | | **PB** | **KrOr** |
| --- | --- | --- | --- | --- | --- | --- | --- | --- | --- | --- |
| Tube B1 | Kappa | Lambda | CD3 | CD20 | CD19 | CD10 | | CD5 | CD23 | CD45 |
| Tube B2 | CD38 | CD79b | - | CD11c | CD19 | CD103 | | CD43 | HLA-DR | CD45 |
| Tube B3 | Kappa | Lambda | CD3 | CD11c | CD19 | CD103 | | CD25 | HLA-DR | CD45 |

*Supplementary Table S1d*. Antibody color combinations for the three 9-color Erlangen panel. B1 is used as the screening panel, B2 for subtype identification, and B3 is the HCL panel.

| Panel | Merged tubes | Shared markers |
| --- | --- | --- |
| MLL9F (base data) | 1, 2 | FS INT LIN, SS INT LIN, CD19, CD45 |
| MLL5F | 2, 3, 4, 5, 7 | FS INT LIN, SS INT LIN, CD19, CD45 |
| Bonn | 1, 2 | FS INT LIN, SS INT LIN, CD19, CD20 |
| Berlin | 2, 3, 4 | FS INT LIN, SS INT LIN, CD19, CD45 |
| Erlangen | B1, B2 | FS INT LIN, SS INT LIN, CD19, CD45 |

*Supplementary Table 2.* Merge parameters for each data set. The tubes that are merged and the shared markers for each data set are reported here. The tubes to be merged are selected to ensure the most extensive set of overlapping CD markers between all data sets. The merge resulted in a combined FCS file with 18 parameters for each event. The merged FCS for the Berlin panel had two additional markers CD43 and CD3; these were ignored for SOM generation as these are not present in the merged base data set.

a)	b)
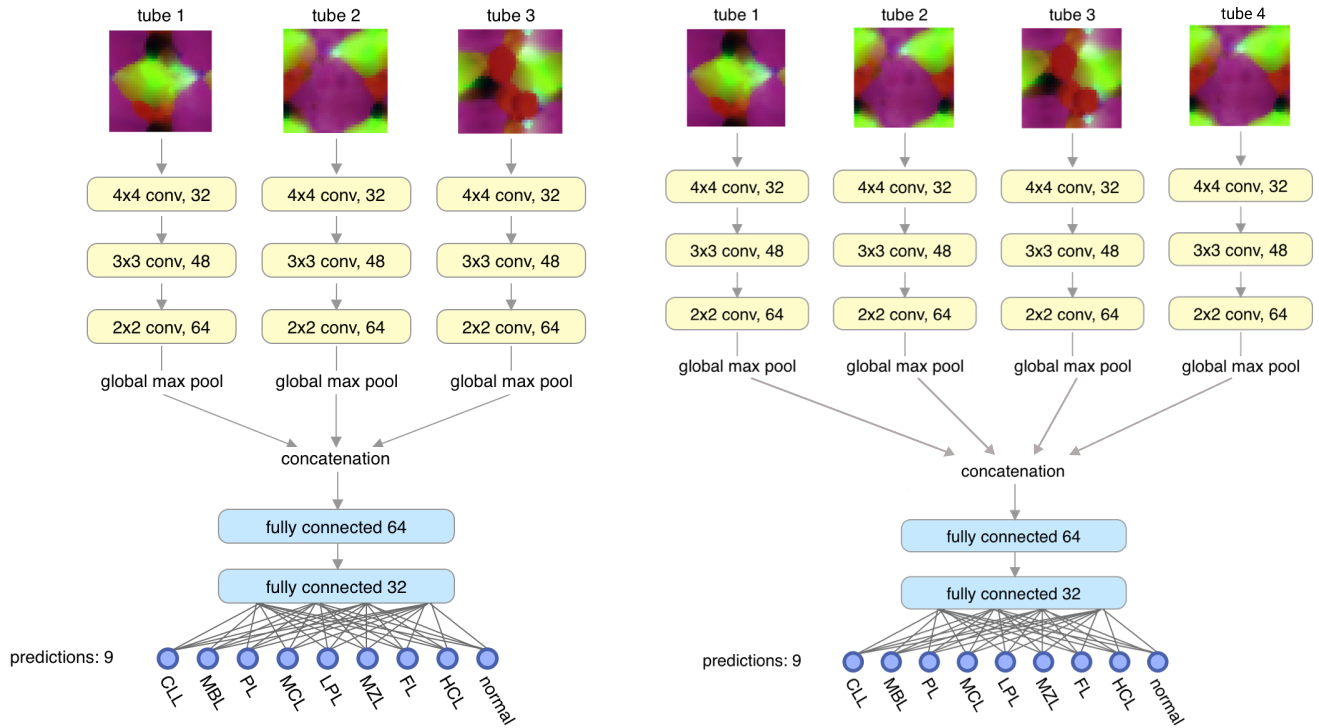

*Supplementary Figure 1.* Original CNN architecture. For each sample, the tubes are processed separately in the convolution layers. There are as many convolution stacks as the number of tubes. When the new data set has a different number of tubes per sample, the number of convolution stacks changes accordingly. a) CNN for a data set with three tubes per sample; three convolution stacks, one per tube. b) CNN for a data set with four tubes is shown. Here, only the weights from dense layers can be transferred between networks. The convolution layers have different data dimensions corresponding to the number of markers per tube and are thus not transferable.

a)
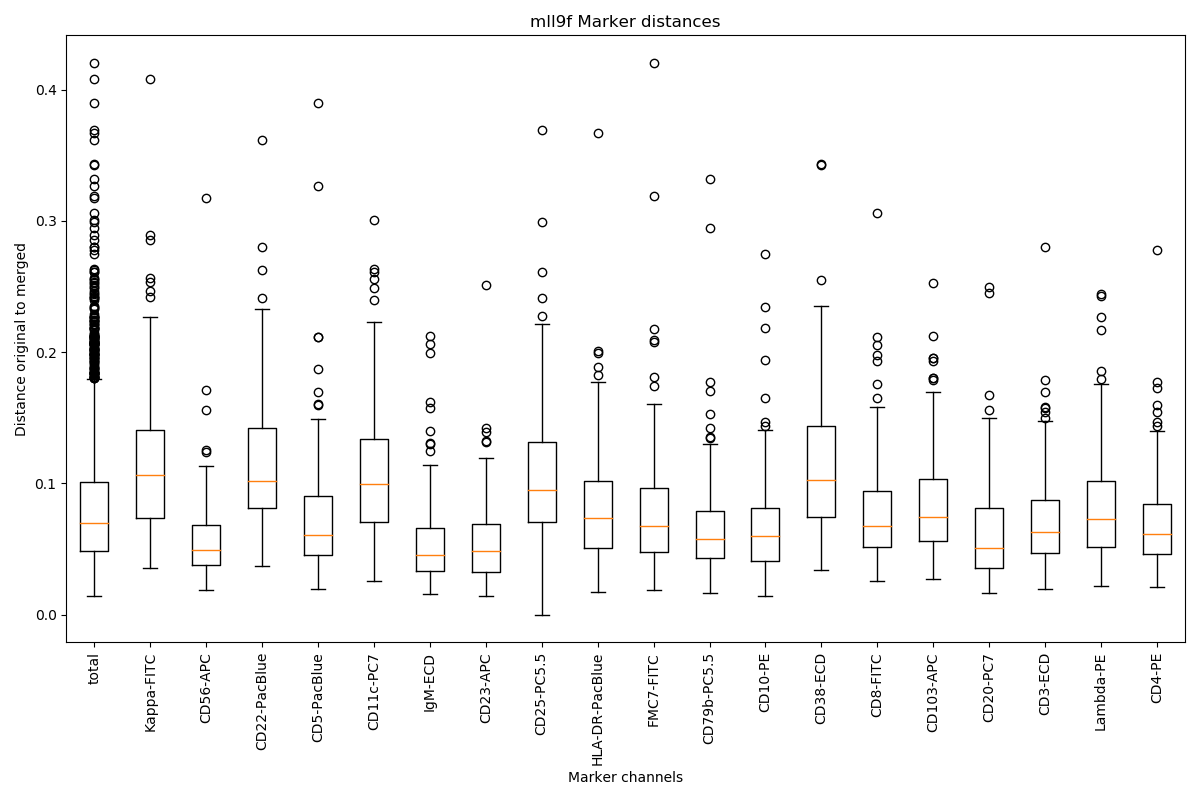
b)

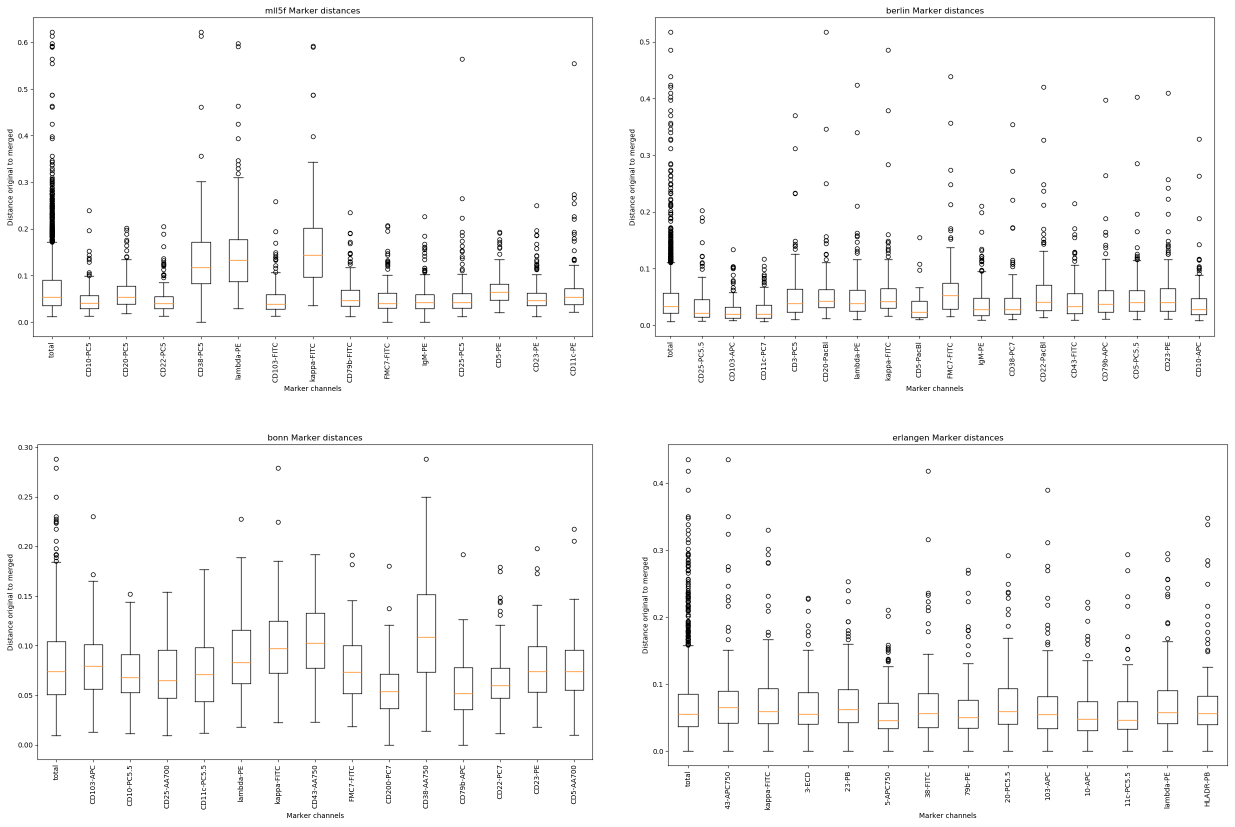

c)
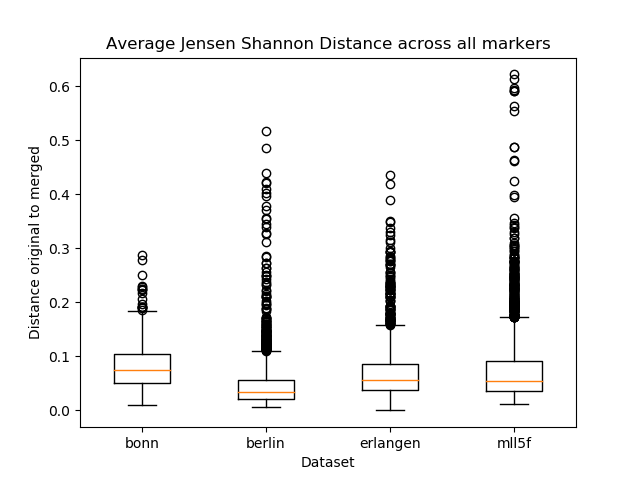

d)

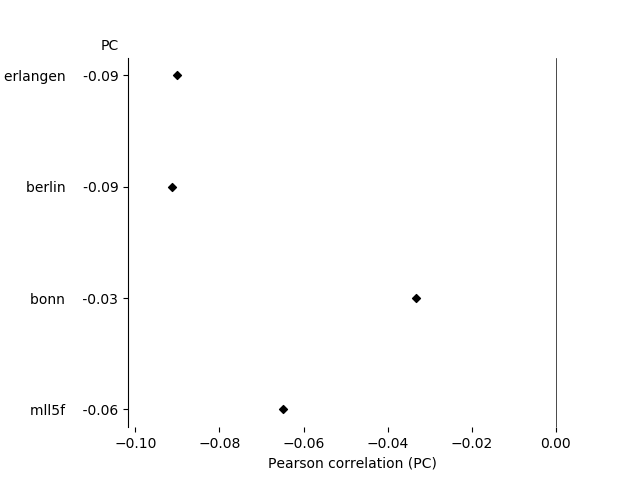

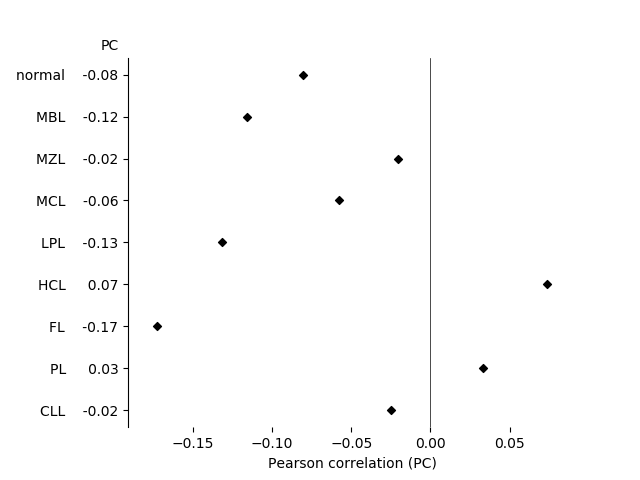

*Supplementary Figure 2.* Jensen-Shannon divergence (JSD) scores for the markers in the base data set are shown in a). The JSD scores for each of the non-shared markers in all the four target data sets are shown in b). The average JSD score across all markers for each target data set is shown in c). We see a mean JSD score of less than 0.1, indicating good agreement between the merged and original data for all four data sets. Fig 2d) shows the correlation between JSD scores and classification accuracy. Pearson correlation (PC) coefficients have been calculated for the true label score in a k-Fold transfer learning experiment for each dataset. Negative PC values show that higher JSD scores correlate with a lower true class prediction score. PC for each dataset shows that all correlations are slightly negative; each group's values also show mostly negative PC scores, except for HCL and PL. Negative PC coefficients indicate that the higher the agreement between merged and original data (low JSD), the higher the classification score.

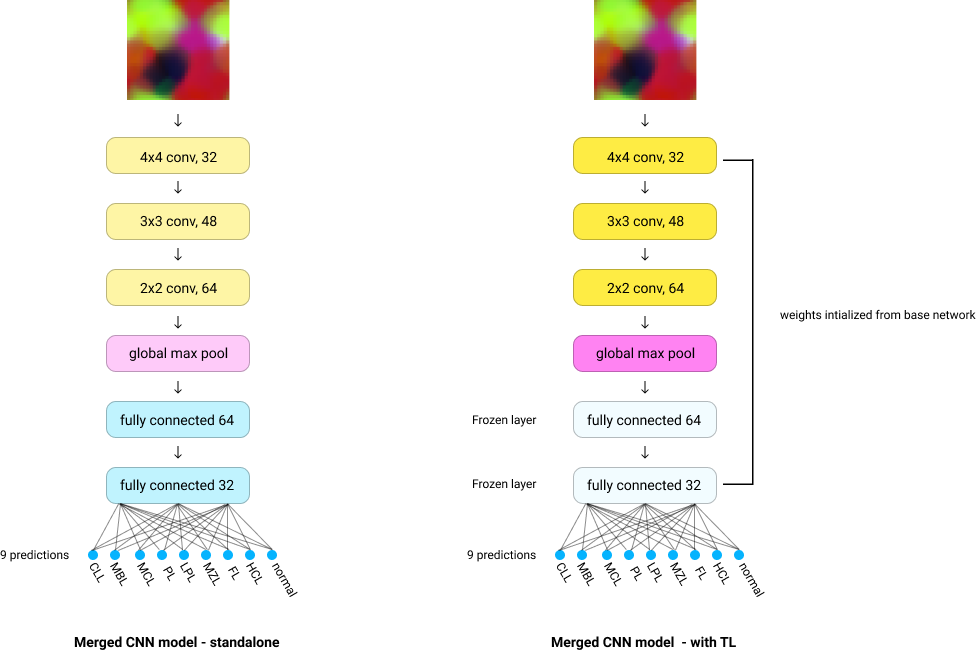
*Supplementary Figure 3*. Modified CNN architecture for the merged data. SOM generated for the merged FCS sample is processed by three convolution layers with varying filter sizes. A single global max pool layer is used after the convolution layers, followed by two fully connected layers that combine the features and generate class predictions. The CNN model with transfer learning has the same parameters as the standalone model; each layer is initialized with weights from the base model. The two dense layers are frozen and not retrained to avoid overfitting.

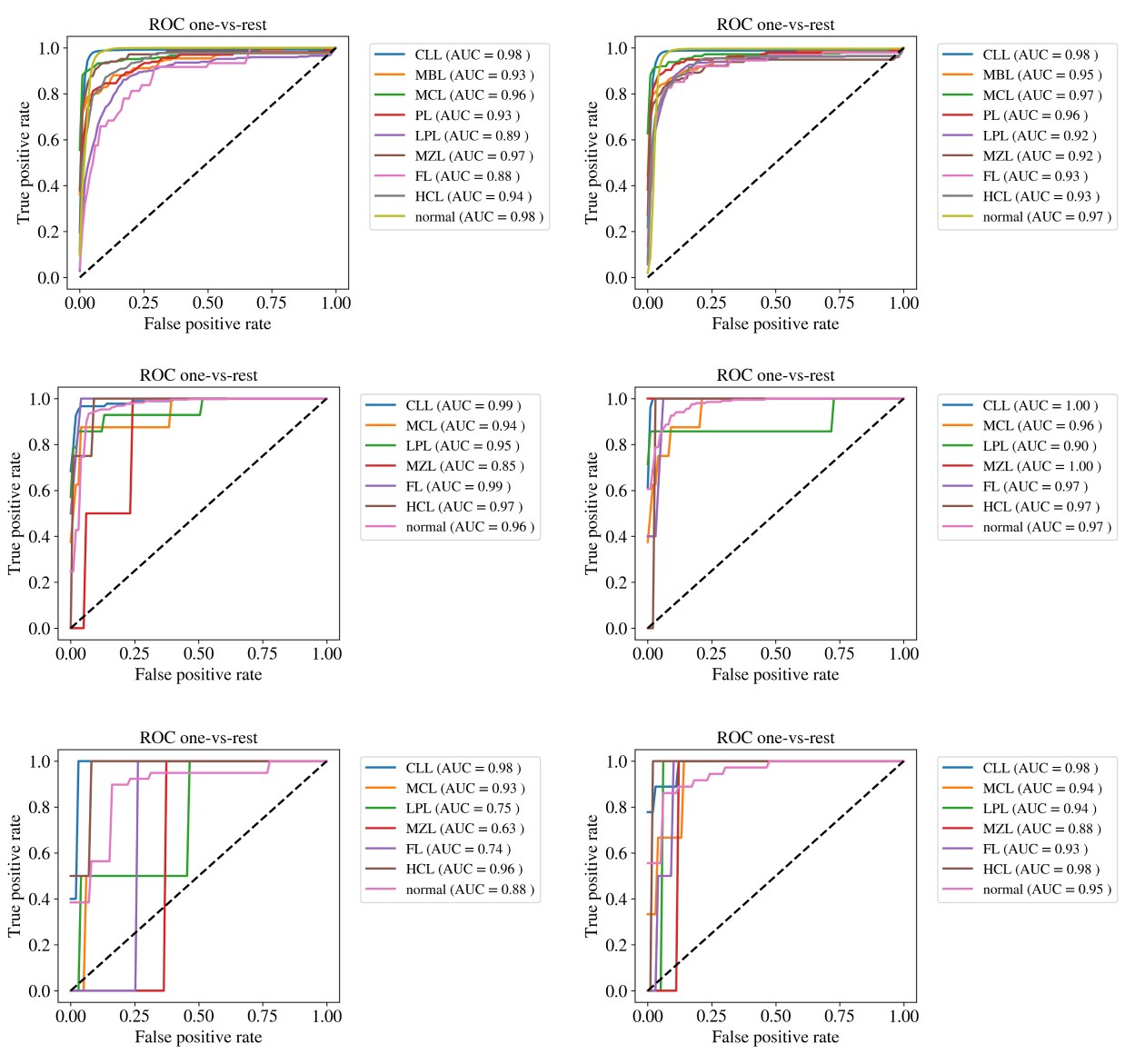

*Supplementary Figure 4*. ROC curves for each data set. The mean ROC curves and AUC for each class across the ten folds are generated in a one-vs-rest manner. Since the data sets are highly imbalanced, there were no validation samples for some of the classes in some folds for Bonn and Berlin panels. Such folds were not used to compute the mean ROC. The first column shows the ROC curves for standalone models, ROC curves with transfer learning are shown in the second column. ROC curves for the MLL5F panel are in the first row, followed by the Berlin panel in the second row. The third row above is for the Bonn panel. As seen in all the rows, the models with transfer learning achieve a higher AUC.
